# Clinical impact of the rapid molecular detection of RSV and influenza A and B viruses in the emergency department

**DOI:** 10.1101/2021.11.18.21264071

**Authors:** Nicolas Yin, Marc Van Nuffelen, Magali Bartiaux, Thierry Préseau, Inge Roggen, Sabrina Delaunoy, Bhavna Mahadeb, Hafid Dahma, Laurent Busson, Olivier Vandenberg, Marie Hallin

## Abstract

**Introduction:** Using respiratory viruses’ rapid diagnostic tests in the emergency department could allow better and faster clinical management. Point-of-care PCR instruments now provide results in less than 30 minutes. The objective of this study was to assess the impact of the use of a rapid molecular diagnostic test, the cobas^®^ Influenza A/B & RSV Assay for use on Roche’s cobas^®^ Liat^®^ instrument, during the clinical management of emergency department patients.

**Methods:** Patients (adults and children) requiring admission or suffering from an underlying condition at risk of respiratory complication were prospectively recruited in the emergency department of four hospitals in the Brussels region. Physician’s intentions regarding admission, isolation, antibiotic, and antiviral use were collected before and after performing the rapid molecular test. Additionally, a comparison of the analytical performance of this test against antigen rapid tests and viral culture was performed as well as a time-to-result evaluation.

**Results:** Among 293 patients recruited, 90 had a positive PCR whereas 44 had a positive antigen test. PCR yielded a sensitivity of 100% for all targets. Antigen tests yielded sensitivities ranging from 66.7% for influenza B to 83.3% for the respiratory syncytial virus (RSV). The use of PCR allowed a decrease in the overall need for isolation and treatment by limiting the isolation of negative patients and the antibiotic use for positive patients. Meanwhile, antiviral treatments better targeted patients with a positive influenza PCR.

**Conclusion:** The use of a rapid influenza and RSV molecular test improves the clinical management of patients admitted to the emergency department by providing a fast and reliable result. Their additional cost compared to antigen tests should be balanced with the benefit of their analytical performance, leading to efficient reductions in the need of isolation and antibiotic use.

## Introduction

The COVID-19 pandemic highlighted the need for rapid and trustworthy diagnostic tests for respiratory tract infections, to assess patients’ potential infectiousness and to set containment measures [1,2]. Various authors have previously demonstrated that a fast and reliable diagnosis of respiratory infections improved the clinical management during the seasonal epidemics of respiratory syncytial virus (RSV) and influenza A and B viruses [3,4]. In Europe, before the COVID-19 pandemic, universal healthcare rarely took the molecular diagnostic of these infections in charge, limiting their prescription in favor of less expensive antigen tests. In Belgium for instance, SARS-CoV-2 PCR detection is now reimbursed, but for RSV and Influenza A and B viruses only antigen detection is [5]. However, compared to RT-PCR, the sensitivity of these antigen rapid diagnostic tests was estimated to be as low as 53.9% in adults and 64.6% in children regarding influenza A [6] and 74% in children regarding RSV [7]; impairing the clinical value of a negative result considerably. Furthermore, clinical judgment, followed by PCR or point-of-care testing (POCT), were found to be cost-effective in a setting where influenza probability was high [8]. Fast “sample-in, result-out” PCR instruments such as the Roche cobas^®^ Liat^®^ are now providing point-of-care PCR results in less than 30 minutes (a run on the instrument is completed in 21 minutes once the sample is loaded; the hands-on time is approximately 5 minutes) [9]. The objective of this study is to assess the impact of the use of the cobas^®^ Liat^®^ Influenza A/B & RSV assay on the clinical management of emergency department (ED) patients requiring admission or presenting an underlying condition at risk of respiratory complication.

## Material and Methods

### Population and data collection

Patients attending the ED of four hospitals located in the Brussels area of Belgium were prospectively recruited. Inclusion criteria were either a pre-test indication of hospitalization or an underlying situation at risk of respiratory complication following influenza infection as described by the European Centre for Disease Prevention and Control and the US Centers for Disease Control (age ≥ 65 years old or < 2 years old, pregnancy, chronic medical conditions, immunocompromised patient, etc.) [10,11]. When including the patient and prescribing the test, physicians indicated their intentions regarding antimicrobial treatment, isolation, and hospitalization by answering a questionnaire. The same questionnaire was completed again once the PCR results were available to evaluate the post-test intentions. The informed consent of each participant or their guardian was collected. The ethics’ committee of each hospital approved the study.

### Specimen collection and analyses

Accepted samples were nasopharyngeal swabs in 3mL of universal transport medium (UTM). Nasopharyngeal aspirates diluted with 3 mL of UTM were also accepted for children. Approximately 200µL were immediately used to perform a rapid PCR using the cobas^®^ Liat^®^ Influenza A/B & RSV assay (Roche Diagnostics, Indianapolis, IN) following manufacturer’s instructions in a 24/7 on-site laboratory or in a point-of-care setting. PCR results were instantly transmitted towards the result servers. PCR time-to-result was assessed for PCR realized in on-site laboratories by the delay between the prescription time indicated by the physician and the results validation time, which is the time it became available for the physician. In addition, antigen rapid diagnostic tests (RDT) were performed after the PCR using Influ A+B K-Set (Coris BioConcept) for Influenza A and Influenza B detection, and RSV K-SeT (Coris BioConcept) for RSV detection on the same sample (200µL/test). Upon arrival in the virology laboratory, 1 mL of UTM was also inoculated on confluent Vero (African green monkey kidney), MRC5 (human lung) and LLC-MK2 (rhesus monkey kidney) cell cultures (Vircell) in 24-well or 6-well tissue culture plates (Greiner-Bio One). Cultures were incubated at 36°C in a 5% CO_2_ atmosphere for 2 weeks for the Vero and LLC-MK2 cells and 3 weeks for the MRC5 cells. The media were replaced weekly. Cultures were examined every two to three days using an inverted microscope. Hemadsorption was performed on the LLC-MK2 cells at the end of the second week of incubation.

### Statistical analysis

To assess the analytical performances of a molecular detection technique against culture and antigen detection, a composite reference standard was constructed as recommended [12]. Samples considered as positive for a viral pathogen were defined as those testing positive for this viral pathogen by at least 2 of the 3 techniques used, negative as those that tested negative by at least 2 of the 3 techniques. Statistical analyses were performed using Analyse-it^®^ for Microsoft Excel v5.30.4. Proportion variations between pre-test and post-test clinical intentions were evaluated using the McNemar-Mosteller exact test.

## Results

Two hundred ninety-three patients were recruited including 68 children (< 15 years old) (Table 1). Among them, 71 had a positive rapid PCR result for influenza A only, 10 for influenza B only, 1 for both influenza A and B, 8 for RSV and 203 were negative for all the targets. The positive agreement between PCR and RDT ranged from 36.3% (4/11) for influenza B to 62.5% (5/8) for RSV. One sample was positive for influenza A using RDT although negative using culture and PCR. Likewise, one sample was positive for influenza A using culture although negative using PCR and RDT. Thus, using the composite reference standard, PCR reached a sensitivity of 100% for the 3 viruses targeted (Table 2).

**Table 1:** Population characteristics

| Age (years) | Female | Male | Overall |
| --- | --- | --- | --- |
| Overall | 123 | 170 | 293 |
| < 15 | 25 | 43 | 68 |
| 15-65 | 63 | 63 | 126 |
| > 65 | 35 | 64 | 99 |

**Table 2:**
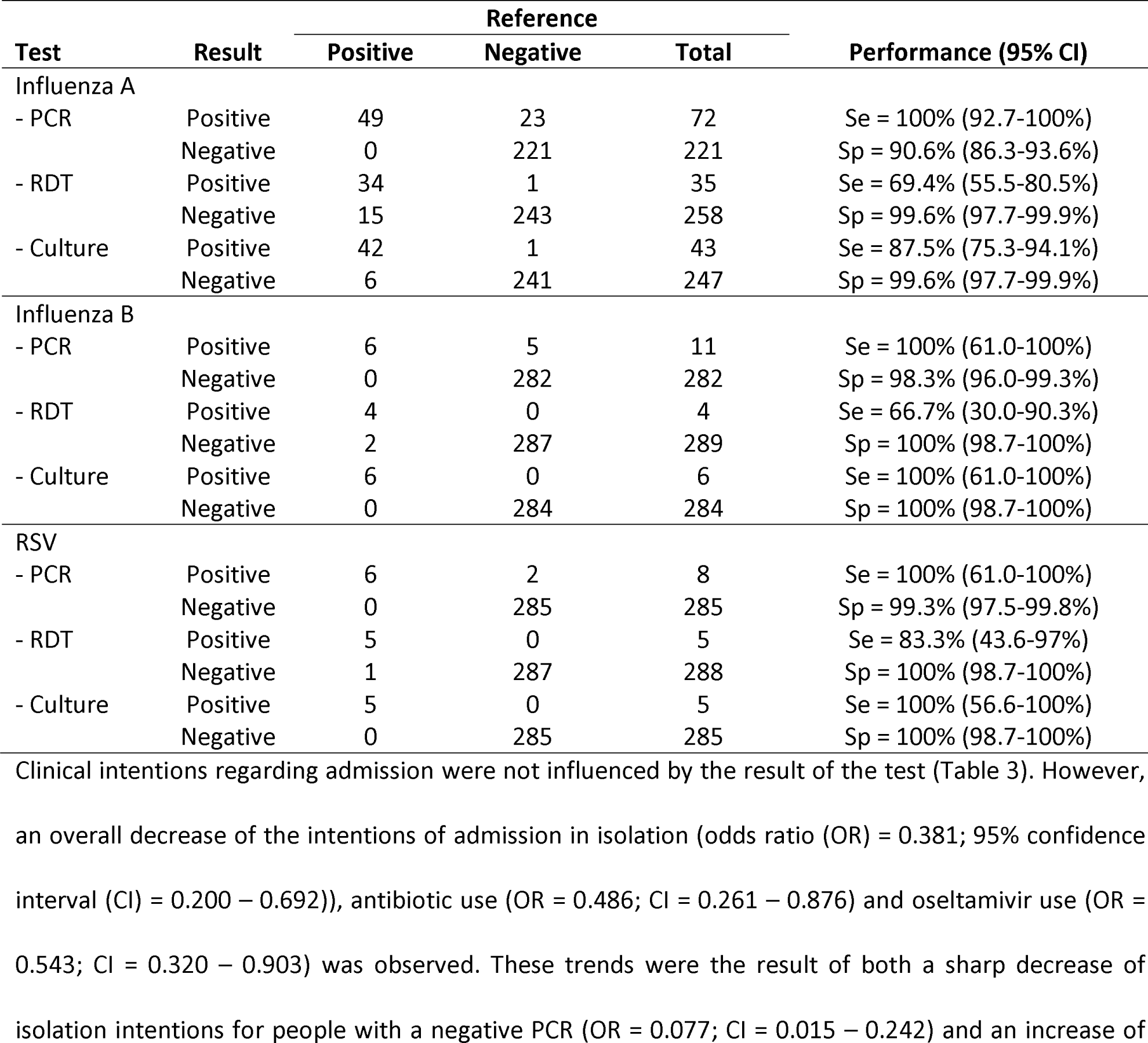
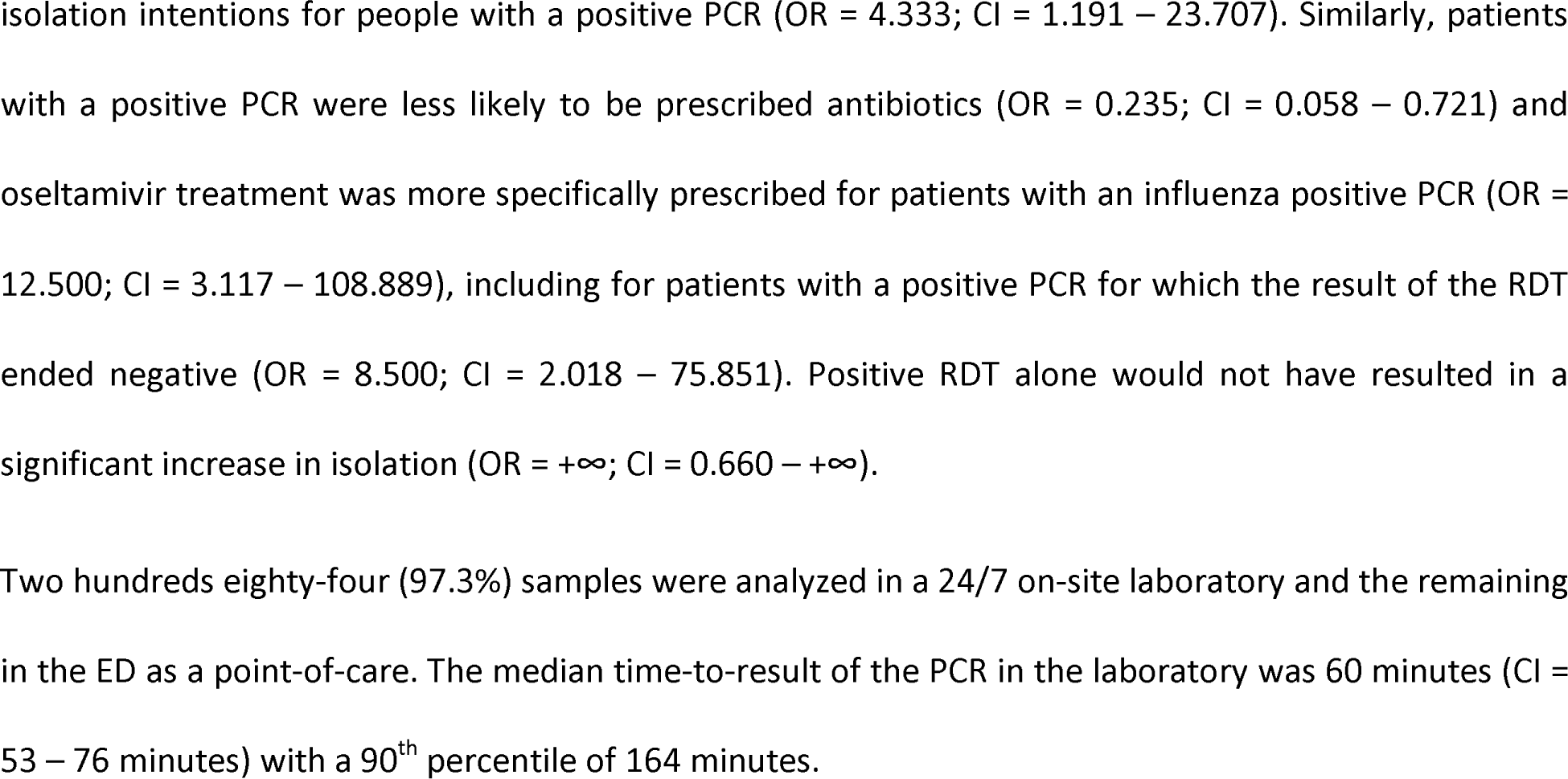
Analytical performance (Se: Sensitivity, Sp: Specificity) and Wilson 95% confidence interval (CI) of Cobas^®^ Liat Influenza A/B & RSV assay (PCR), antigen tests (RDT) and viral culture for the diagnostic of Influenza A, B and RSV using a composite reference standard

**Table 3:**
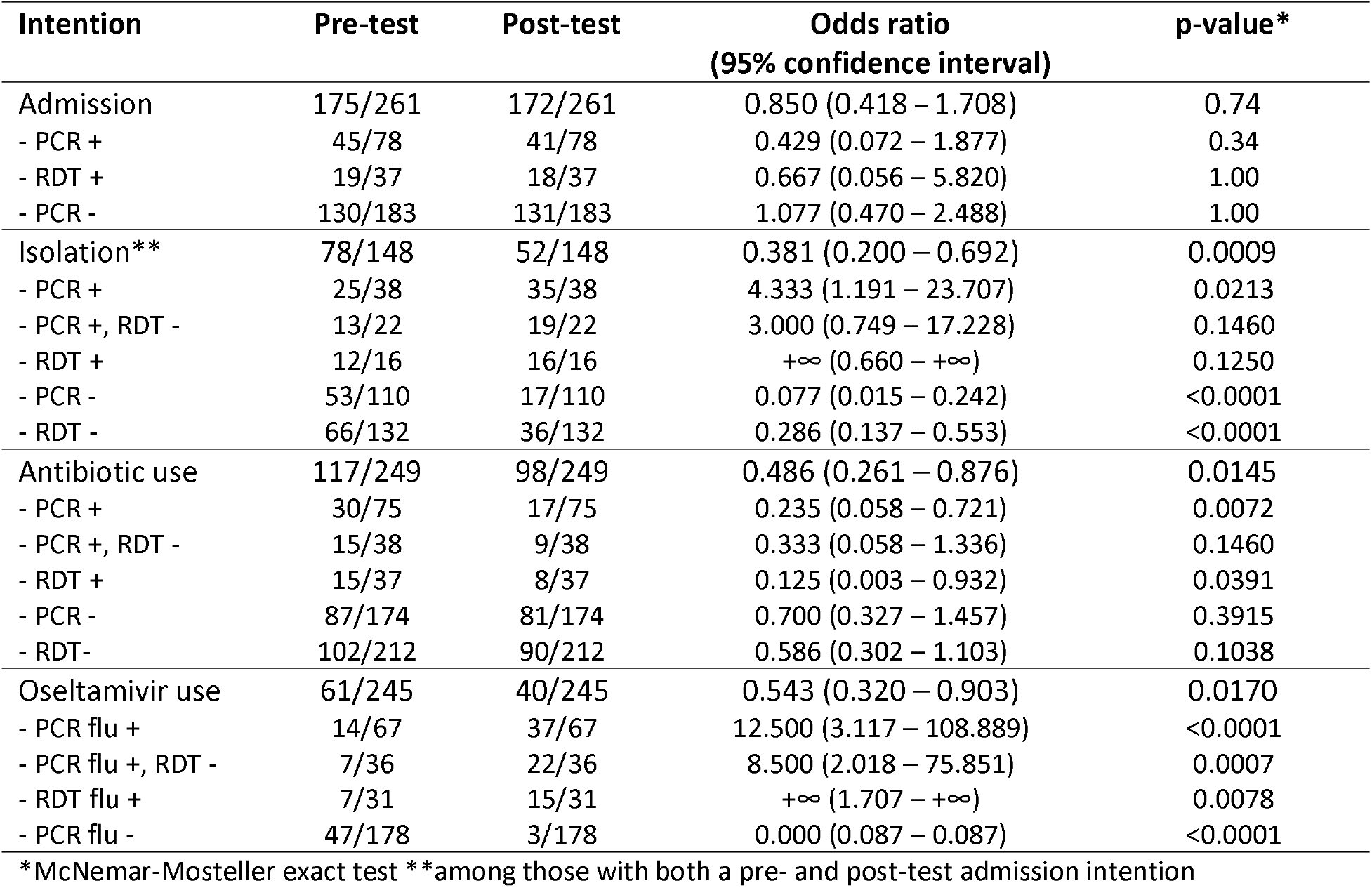
Clinical impact of the positive (+) and negative (-) results of the PCR and antigen rapid diagnostic test (RDT) for respiratory syncytial virus (RSV) and influenza A and B viruses (flu)

## Discussion

Previous studies already underlined the impact of the rapid detection of influenza and RSV in the management of patients [13–16]. We showed that an overall significant decrease of antibiotic prescription could be obtained by providing a relevant diagnostic tool combining speed and sensitivity. The excellent sensitivity of the cobas^®^ Liat^®^ assay allowed an optimized assessment of the need for isolation for the inpatients, ensuring a proper isolation for almost all infected patients, which likely led to a reduction of nosocomial transmission. Indeed, less than half of patients with a positive PCR had also a positive RDT underlining the importance to use a molecular test efficiently improving antibiotic and antiviral stewardship while offering a real assessment for the need of a costly hospitalization in isolation. The overall decrease of the isolation indications for patients with a negative test should be balanced with the clinical presentation of the patient to prevent the nosocomial transmission of eventual other respiratory pathogens. Nevertheless, in a pandemic context where isolation resources can be scarce, the use of a negative reliable test to decrease the need for isolation should be considered.

In our study, the median time of response from the laboratory was 60 minutes for a less than 30-minute assay. Nevertheless, the time-to-result in this setting is dependent of the overall workload in the on-site laboratory. Using this assay as a POCT at the ED would certainly decrease the time-to-result while increasing the autonomy of the ED team regarding the general management of the workflow and the real-time adaptation of the indications depending on the current level of bed occupancy and the epidemic situation.

However, such a strategy has a cost and should be further examined through a medico-economic study to better determine the optimal target population, while taking into account the subsequent decrease of nosocomial outbreaks and the targeted use of individual protection equipment and antibiotics. In a Dutch study, the daily direct cost of isolating patients ranged from €28 (£23/$31) to €41 (£34/$46) [17]. At the time of writing, the price of the cobas^®^ Liat^®^ Influenza A/B & RSV was 39€/test (£33/$44) (excluding instruments and workforce). In Belgium, an antigen test is reimbursed 8.06€ (£6.74/$9.00) per target (maximum 3) and a viral culture, 45.17€ (£37.75/$50.48). The savings will likely balance the costs of performing a PCR instead of RDTs and viral cultures for inpatients during seasonal epidemic.

Even if we were able to evaluate the cobas^®^ Liat^®^ Influenza A/B & RSV assay in the laboratory, this study was prematurely interrupted by the surge of the COVID-19 pandemic in Belgium in March 2020. This event prevented us to evaluate the benefit of the use of this assay as a POCT because this assay was not anymore relevant for the epidemic at the time. The need for human and technical resources also prevented us to cross check PCR-only positive tests with another molecular method leading here to a likely underestimation of the specificity and the need to use a composite reference standard to balance the well-known lesser sensitivity of RDT and culture. It also restricted the number of RSV and influenza B infections. However, the pandemic allowed us to later use the cobas^®^ Liat^®^ SARS-CoV-2 & Influenza A/B assay as a POCT in the ED. This implementation raised biosafety concerns, as this assay requires the transfer by pipetting of the transport medium to the reagent tube, which can be considered as a projection and lead to aerosolization risks. To solve this, the loading of the reagent tube with the UTM was done at bedside, by the operator performing the sampling wearing personal protection equipment. The sealed tube was then transported to the instrument located in a separate room [18].

In conclusion, using a rapid molecular assay, such as the cobas^®^ Liat^®^ Influenza A/B & RSV, improves the clinical management of patients by refining the indications of isolation, antibiotic, and antiviral treatment thanks to better analytical performances than RDTs. Using this instrument as a POCT in the ED could provide an efficient 24/7 solution to dispatch inpatients and take at-risk outpatients in charge swiftly and optimally.

## Data Availability

The raw data supporting the conclusions of this article will be made available by the authors, without undue reservation.

## Acknowledgment

We wish to thank the nurses of the emergency department of our hospitals for their amazing work and their collaboration in this study. We would like to thank Dustin DeMeglio for his help in English writing.

## References

1. Vandenberg O, Martiny D, Rochas O, van Belkum A, Kozlakidis Z. Considerations for diagnostic COVID-19 tests. Nat Rev Microbiol. 2021 Mar;19(3):171–83.

2. Sheridan C. Coronavirus testing finally gathers speed. Nat Biotechnol [Internet]. 2020 Nov 5 [cited 2020 Dec 20]; Available from: http://www.nature.com/articles/d41587-020-00021-z

3. Maltezou HC. Nosocomial influenza: new concepts and practice: Curr Opin Infect Dis. 2008 Aug;21(4):337–43.

4. Bont L. Nosocomial RSV infection control and outbreak management. Paediatr Respir Rev. 2009 Jun;10:16–7.

5. Nomenclature des prestations de santé [Internet]. Sect. Biologie Clinique - Article 24. Available from: https://www.inami.fgov.be/SiteCollectionDocuments/nomenclatureart24_20210701_01.pdf

6. Chartrand C, Leeflang MMG, Minion J, Brewer T, Pai M. Accuracy of Rapid Influenza Diagnostic Tests: A Meta-analysis. Ann Intern Med. 2012 Apr 3;156(7):500.

7. Chartrand C, Tremblay N, Renaud C, Papenburg J. Diagnostic Accuracy of Rapid Antigen Detection Tests for Respiratory Syncytial Virus Infection: Systematic Review and Meta-analysis. Tang Y-W, editor. J Clin Microbiol. 2015 Dec;53(12):3738–49.

8. Lee BY, McGlone SM, Bailey RR, Wiringa AE, Zimmer SM, Smith KJ, et al. To Test or to Treat? An Analysis of Influenza Testing and Antiviral Treatment Strategies Using Economic Computer Modeling. Galvani AP, editor. PLoS ONE. 2010 Jun 23;5(6):e11284.

9. Gibson J, Schechter-Perkins EM, Mitchell P, Mace S, Tian Y, Williams K, et al. Multi-center evaluation of the cobas ® Liat ® Influenza A/B & RSV assay for rapid point of care diagnosis. J Clin Virol. 2017 Oct;95:5–9.

10. European Centre for Disease Prevention and Control. Factsheet about seasonal influenza [Internet]. [cited 2021 Jul 27]. Available from: https://www.ecdc.europa.eu/en/seasonal-influenza/facts/factsheet

11. Centers for Disease Control and Prevention. Influenza Antiviral Medications: Summary for Clinicians [Internet]. [cited 2021 Jul 27]. Available from: https://www.cdc.gov/flu/professionals/antivirals/summary-clinicians.htm

12. Banoo S, Bell D, Bossuyt P, Herring A, Mabey D, Poole F, et al. Evaluation of diagnostic tests for infectious diseases: general principles. Nat Rev Microbiol. 2010 Dec;8(S12):S16–28.

13. Bonner AB, Monroe KW, Talley LI, Klasner AE, Kimberlin DW. Impact of the Rapid Diagnosis of Influenza on Physician Decision-Making and Patient Management in the Pediatric Emergency Department: Results of a Randomized, Prospective, Controlled Trial. PEDIATRICS. 2003 Aug 1;112(2):363–7.

14. Rahamat-Langendoen J, Groenewoud H, Kuijpers J, Melchers WJG, Wilt GJ. Impact of molecular point-of-care testing on clinical management and in-hospital costs of patients suspected of influenza or RSV infection: a modeling study. J Med Virol. 2019 Aug;91(8):1408–14.

15. Falsey AR, Murata Y, Walsh EE. Impact of rapid diagnosis on management of adults hospitalized with influenza. Arch Intern Med. 2007 Feb 26;167(4):354–60.

16. Busson L, Bartiaux M, Brahim S, Konopnicki D, Dauby N, Gérard M, et al. Contribution of the FilmArray Respiratory Panel in the management of adult and pediatric patients attending the emergency room during 2015–2016 influenza epidemics: An interventional study. Int J Infect Dis. 2019 Jun;83:32–9.

17. van Dijk MD, Voor in ‘t holt AF, Polinder S, Severin JA, Vos MC. The daily direct costs of isolating patients identified with highly resistant micro-organisms in a non-outbreak setting. J Hosp Infect. 2021 Mar;109:88–95.

18. Yin N, Debuysschere C, Decroly M, Bouazza F-Z, Collot V, Martin C, et al. SARS-CoV-2 Diagnostic Tests: Algorithm and Field Evaluation From the Near Patient Testing to the Automated Diagnostic Platform. Front Med. 2021;8:650581.

